# Behavioral and neural dissociation of social anxiety and loneliness

**DOI:** 10.1101/2021.08.25.21262544

**Authors:** Jana Lieberz, Simone G. Shamay-Tsoory, Nira Saporta, Alisa Kanterman, Jessica Gorni, Timo Esser, Ekaterina Kuskova, Johannes Schultz, René Hurlemann, Dirk Scheele

## Abstract

**Background:** Loneliness is a public health concern with detrimental effects on physical and mental well-being. Given phenotypical overlaps between loneliness and social anxiety, cognitive behavioral interventions targeting social anxiety might be adopted to reduce loneliness. However, it is still elusive whether social anxiety and loneliness share the same underlying neurocognitive mechanisms. The current study aimed at investigating to what extent known behavioral and neural correlates of social avoidance in social anxiety are evident in loneliness.

**Methods:** We used a pre-stratified approach involving 42 participants with high and 40 control participants with low loneliness scores. During functional magnetic resonance imaging, the participants completed a social gambling task to measure the subjective value of engaging in a social situation and responses to positive and negative social feedback.

**Results:** Uni- and multivariate analyses of behavioral and neural data replicated known task effects across groups. However, although lonely participants were characterized by increased social anxiety, loneliness was associated with a response pattern clearly distinct from social anxiety. Specifically, Bayesian analyses revealed moderate evidence for equal subjective values of engaging in social situations and comparable amygdala responses to social decision-making and striatal responses to positive social feedback in both groups. Conversely, lonely participants showed significantly altered behavioral responsiveness to negative feedback and reduced striatal activity, whereas striatal-hippocampal connectivity was increased compared to controls.

**Conclusion:** Our findings suggest that loneliness is associated with altered emotional reactivity to social situations rather than behavioral tendencies to withdraw from social interactions. Thus, established interventions for social anxiety should be adjusted when targeting loneliness.

## Introduction

Loneliness is a painful condition which can be a catalyst for subjective stress (1) and is associated with detrimental effects on mental and physical health (2, 3). As such, loneliness has been identified as a risk factor for premature mortality comparable with smoking or obesity (4, 5). Consequently, loneliness has come into focus of politics and clinicians as a major public health concern with high economic costs for society (6-8). With social distancing in place in most countries around the world, COVID-19 is expected to have vast impact on physical and mental health, particularly in people inflicted by poor resilience to social adversity due to pre-existing low levels of social integration (9, 10). Preliminary evidence indicate that the prevalence of loneliness might have increased due to the ongoing COVID-19 pandemic, which emphasizes the urgent need of interventions to target loneliness (11-14).

Recent findings highlight a close link of loneliness with social anxiety symptoms (15-17) and identified social anxiety as predictor for future loneliness (18-20). For instance, social anxiety was found to be consistently associated with social isolation, lower perceived social support, and decreased relationship satisfaction (21-23). Moreover, poor friendship quality promotes increases in social anxiety symptomatology (24). A perceived discrepancy in the quality and quantity of the actual and desired relationships, in turn, is a key feature of loneliness (25). Likewise, safety behavior such as the avoidance of social situations is known to be a core mechanism fostering the maintenance of social anxiety and is also hypothesized to be preferred by lonely individuals (26, 27).

Given the phenotypical overlap between loneliness and social anxiety, cognitive behavioral therapies targeting social anxiety might be co-opted as interventions to reduce loneliness. Existing programs are often based on cognitive models of social anxiety (28), which posit an exaggerated fear of evaluation as a core etiological mechanism of psychopathology. Indeed, current neurocircuitry models of social anxiety disorder emphasize amygdala hyperreactivity to social stimuli (29, 30) and we have recently observed increased amygdala responses during social decision-making and social feedback in healthy individuals with high social anxiety (31). By contrast, the neural responsiveness to social rewards such as happy faces seems to be reduced in individuals with social anxiety (31-34), potentially resulting in reduced positive affect in response to social interactions and impaired memory for positive social events (35, 36). Similarly, lonely individuals exhibit an attenuated responsiveness to positive social interactions (37) and there is preliminary evidence indicating that alterations in amygdala structure and function are associated with loneliness (for a current comprehensive review of neurobiological factors associated with loneliness, see (38)). However, it is still elusive whether social anxiety and loneliness share similar neurobiological substrates during social interactions or whether psychotherapeutic protocols need to be adjusted to reduce chronic loneliness.

The current study therefore aims at examining whether previously reported mechanisms underlying social anxiety (cf. (31)) could be replicated in loneliness. Thus, we recruited a pre-stratified sample of 42 healthy participants scoring high (high-lonely, HL) and 40 control participants scoring low (low-lonely, LL) on a loneliness scale. During functional magnetic resonance imaging (fMRI), the participants completed a social gambling task to measure the subjective value of engaging in a social situation and responses to positive and negative social feedback. The task has been previously used to identify a potential neural circuitry underlying the social avoidance behavior associated with social anxiety (cf. (31)). Given the intertwined phenotype of both constructs, we hypothesized that lonely individuals would exhibit a decreased subjective value of engaging in social situations as observed for social anxiety. Likewise, we expected increased amygdala activation during social decision-making and social feedback and concomitantly decreased reward-associated responses of the nucleus accumbens (NAcc) to positive social feedback in lonely participants. We further tested whether changes in brain activity were associated with altered functional connectivity. In an additional exploratory analysis, we examined distinct behavioral and neural response patterns in loneliness that have not been previously found to be associated with social anxiety (i.e., responsiveness to negative social feedback). Notably, we controlled for the influence of social anxiety and further potential confounding variables including depressive symptomatology and childhood maltreatment for all observed associations of loneliness with neural or behavioral measurements.

## Materials and Methods

### Participants

We recruited a sample of 82 (out of a stratified sample of 3678 adults; 41 females, mean age ± standard deviation (SD): 26.83 ± 7.47 years) pre-stratified healthy volunteers with high (*n* = 42) and low loneliness scores (*n* = 40) as assessed by the revised version of the UCLA loneliness scale (UCLA-L, (39); for details, see supplementary material and (37)). All participants gave written informed consent. The study was approved by the institutional review board of the Medical Faculty of the University of Bonn (study number 016/18) and conducted in accordance with the latest revision of the Declaration of Helsinki.

### Behavioral tasks

We measured the participants’ subjective value of engaging in social situations with a social gambling task (cf. (31) and see supplementary material). During a decision phase, participants could choose a risky (a dice game with a virtual human or computer partner with equiprobable outcomes of 3 or 0 €) or a safe option (a fixed payoff ranging from 0 to 3 €). If participants chose the risky option, either a positive or a negative feedback video of the partner (human or computer) was shown (feedback phase), depending on the outcome of the trial (win or loss). As such, the human feedback video displayed the virtual human partner expressing either admiration or condescension. All human videos were taken from a validated database (40). In the computer control condition, the feedback was given by a video of a green checkmark (participant won) or a red cross (participant lost). If participants chose the safe option, a sentence confirmed the payoff. Individual certainty equivalents of the risky option (termed CE50), i.e., the certain payoff for which a participant would be indifferent between the risky and safe options (i.e., they would choose each option with equal probability), were estimated separately for the computer and the human partners by fitting participants’ choices with a cumulative Gaussian function. CE20 and CE80, i.e., certain payoffs associated with choosing the safe option with 20 % and 80 % probability, were similarly estimated. The subjective value of engaging in social situations was defined as the individual difference between the estimated CE50 for human partners compared to the computer partner. After finishing the task, the pleasantness of each feedback video was rated on a visual analogue scale ranging from 0 (“not pleasant at all”) to 100 (“very pleasant”). The task was then repeated during functional magnetic resonance imaging (fMRI) with randomly chosen partners (human or computer) for each trial. The fixed payoff offered as safe option varied randomly between the individually determined values CE20, CE50, and CE80. Using individualized payoffs as a safe alternative enabled us to equate the number of risky and safe choices across participants.

We further measured the individual monetary value associated with receiving positive or avoiding negative social feedback during a virtual auction task. Specifically, participants were informed that they were participating in a virtual auction against the computer using a random algorithm to invest money. Participants were then asked with no imposed time limit to invest any amount of money between 0 € and 1 € (in increments of 5 cents) to (1) increase the probability of watching a positive social feedback video or (2) to decrease the probability of watching a negative social feedback video (see supplementary material).

### Statistical analyses

Behavioral data were analyzed in SPSS 24 (IBM Corp., Armonk, NY) by calculating analyses of variance (ANOVAs) and Bonferroni-corrected (*P*_cor_) post-hoc t-tests. *P*-values < 0.05 (two-tailed) were considered significant. To analyze the fMRI data, we used a two-stage approach as implemented in SPM12 (Wellcome Trust Center for Neuroimaging, London, UK; http://www.fil.ion.ucl.ac.uk/spm). On the first level, data were modeled using a fixed-effects model. Within-subject contrasts of interest were entered to a random-effects model on the second level to assess group-specific response patterns by calculating two-sample t-tests. Specifically, to probe the hypothesis of increased amygdala activation during social decision-making in HL participants, we compared brain activity during risky decisions involving a human partner between groups by calculating two-sample t-tests (i.e., HL _risky decision human > safe decision human_ > LL _risky decision human > safe decision human_, HL _risky decision human > risky decision computer_ > LL _risky decision human > risky decision computer_). Likewise, the hypothesized increased amygdala responsiveness to social feedback (HL _human feedback > computer feedback_ > LL _human feedback > computer feedback_) and reduced NAcc reactivity to positive social feedback (LL _positive human feedback > positive computer feedback_ > HL _positive human feedback > positive computer feedback_) were tested by calculating two-sample t-tests. As the behavioral data indicated an altered responsiveness to negative social feedback (see results), we explored group differences in response to negative human feedback videos (HL _negative human feedback > negative computer feedback_ > LL _negative human feedback > negative computer feedback_). These contrasts were also calculated in the opposite direction (e.g., LL _risky decision human > risky decision computer_ > HL _risky decision human > risky decision computer_). The amygdala and NAcc were anatomically defined according to the Wake Forest University PickAtlas (41, 42). *P*-values < 0.05 after familywise error correction for multiple testing (*P*_FWE_) based on the size of the respective region of interest were considered significant. Parameter estimates of clusters showing significant group effects were extracted and further analyzed in SPSS 24 to disentangle the group x task condition interaction. Behavioral group effects were correlated with parameter estimates of neural group effects. For details, see supplementary material. The analysis plan was preregistered prior to conducting any analyses (https://osf.io/x47ke). All data used in this study are openly available (https://osf.io/p6jxk/ and https://neurovault.org/collections/VNYRMORR/).

### Explorative analyses

We conducted a multivariate pattern analysis using the Decoding Toolbox (43) to test whether we could replicate previous findings that decisions of the participants could be decoded from amygdala activation (cf. (31)). Contrasts revealing significant group effects in the univariate activity analyses (see above) were further examined by generalized psychophysiological interaction (gPPI) analyses using the CONN toolbox 19.b (www.nitrc.org/projects/conn, RRID:SCR_009550). Mediation and moderation analyses were run to examine the potential influence of depressive and social anxiety symptoms (assessed by the Beck’s Depression Inventory II, BDI (44) and the Liebowitz Social Anxiety Scale, LSAS (45)) and childhood maltreatment (assessed by the Childhood Trauma Questionnaire, CTQ (46)) on observed loneliness effects. For hypotheses that could not be confirmed, we conducted Bayesian t-tests using JASP (47) to quantify the evidence for an absence of group differences. For details of the explorative analyses, see supplementary material.

## Results

## Behavioral results

As expected, social anxiety was significantly increased in HL participants (*t*(67.74) = 3.25, *P* = 0.002, *d* = 0.72; mean LSAS score ± SD in HL: 18.64 ± 15.91; LL: 9.28 ± 9.56; see (37) and **Fig. 1A**) and task effects of the social gambling task reported by (31) were replicated across groups (see supplementary material and **Fig. 1B**). However, contrary to previously observed effects of social anxiety (31), loneliness (HL vs. LL) affected neither the subjective value of engaging in social situations during the behavioral social gambling task nor the invested money in the virtual auction task (all *P*s > 0.05).

**Fig. 1.**
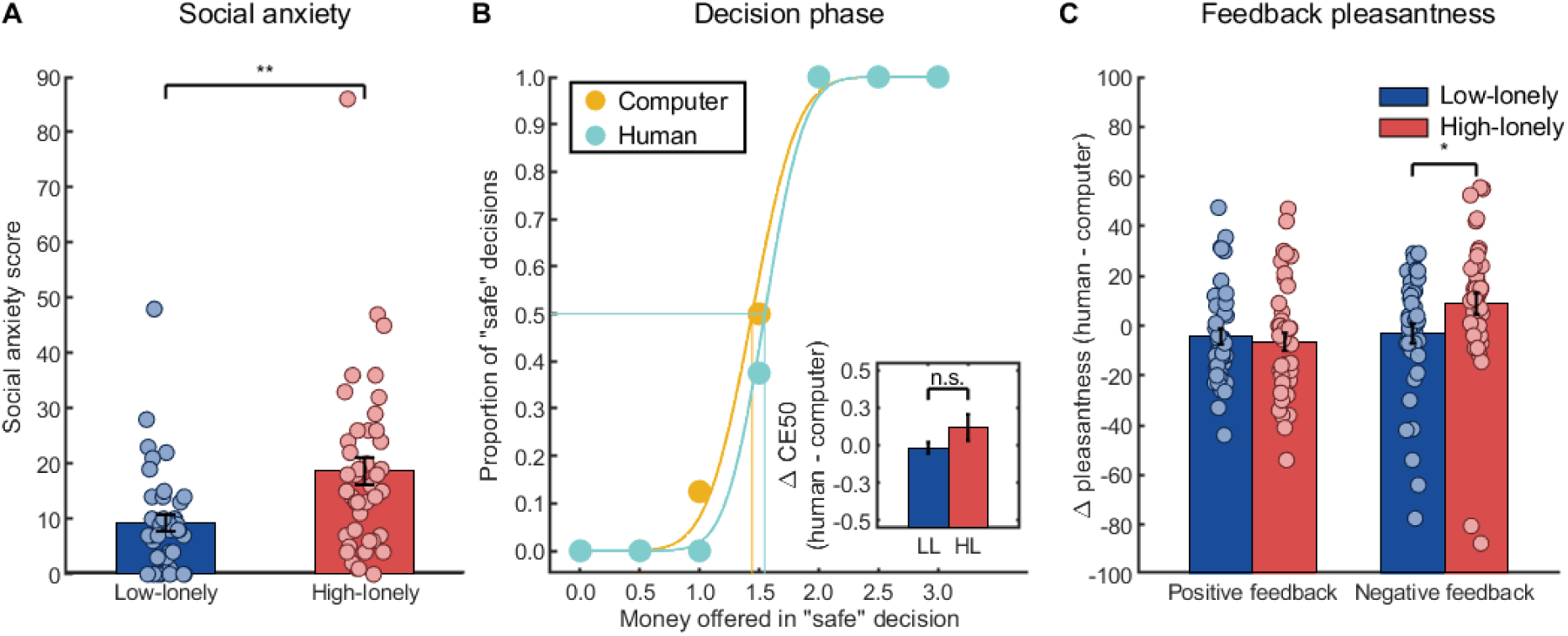
Behavioral results of the decision and feedback phase of the social gambling task. (**A**) Participants with a high loneliness score (HL) showed significantly increased social anxiety scores as assessed with the Liebowitz Social Anxiety Scale. **(B)** The proportion of safe decisions during the social gambling task increased with higher payoffs offered in those safe decisions (main effect of offered payoff for the behavioral task: F(2.95,236.14) = 183.77, P < 0.001, η_p_^2^ = 0.70; functional magnetic resonance imaging task: F(2,158) = 185.43, P < 0.001, η_p_^2^ = 0.70; example data of the behavioral task from one HL participant are presented). As presented in the inlay, HL participants did not significantly differ from control participants (LL) with regard to the subjective value of engaging in a social situation (i.e., CE50, the payoff offered in the safe option associated with 50% of safe decisions). (**C**) By contrast, groups significantly differed in their pleasantness ratings of the negative feedback videos. Compared to the negative computer feedback video, HL participants rated the negative human feedback video as more pleasant, whereas LL control participants showed the opposite pattern of ratings. No differences between groups were observed for positive feedback. Each marker in (B) represents the mean of 8 trials. Bars represent group means. Error bars indicate standard errors of the mean. Abbreviations: n.s., not significant. * P < 0.05, ** P < 0.01.

Nevertheless, analyses of pleasantness ratings of the feedback videos revealed a significant interaction of group x partner x feedback valence (*F*(1,80) = 4.02, *P* = 0.048, η_p_^2^ = 0.05). To disentangle the interaction, we calculated further mixed ANOVAs separately for the positive and negative feedback videos. Surprisingly, no group effects were observed for positive feedback (all *P*s > 0.05), but we found a significant interaction of group x partner for negative feedback (*F*(1,80) = 4.34, *P* = 0.04, η_p_^2^ = 0.05; see **Fig. 1C**). HL participants rated the negative human feedback video as more pleasant compared to the negative computer feedback (*t*(41) = 2.09, *P*_cor_ = 0.09), while LL participants showed the opposite pattern of ratings (*t*(39) = -0.82, *P*_cor_ = 0.84).

### fMRI results

Multi- and univariate analyses of neural activation across groups replicated all previous task effects (31). As such, a linear support vector machine classifier based on amygdala activation was able to decode the decision (risky vs. safe) significantly better than chance (mean accuracy ± SD = 53.64 ± 9.07 %; 30, -4,, 28, *t*(73) = 3.45, *P*_FWE_ = 0.048). Amygdala activation increased during decisions involving a human partner compared to the computer partner (right: 22, -6, -12, *t*(73) = 3.68, *P*_FWE_ = 0.03; left: -22, -8, -12, *t*(73) = 4.00, *P*_FWE_ = 0.01). Specifically, amygdala activity was enhanced during trials in which participants chose the risky option with a human partner compared to the computer partner (right: 22, -6, -12, *t*(73) = 4.58, *P*_FWE_ = 0.002; left: -22, -8, -12, *t*(73) = 4.23, *P*_FWE_ = 0.006; see **Fig. 2A**), while no differences in amygdala activity between partners were observed for safe decisions. Moreover, receiving feedback from the human partner activated the amygdala significantly stronger than computer feedback (right: 22, -6, -14, *t*(75) = 9.67, *P*_FWE_ < 0.001, left: -22, -8, -12, *t*(75) = 9.66, *P*_FWE_ < 0.001) and NAcc activity across partners was increased in response to positive feedback compared to negative feedback (right: 12, 8, -6, *t*(75) = 6.45, *P*_FWE_ < 0.001, left: -14, 10 -10, *t*(75) = 4.91, *P*_FWE_ < 0.001). For whole-brain task effects, see **Table S1**.

**Fig. 2.**
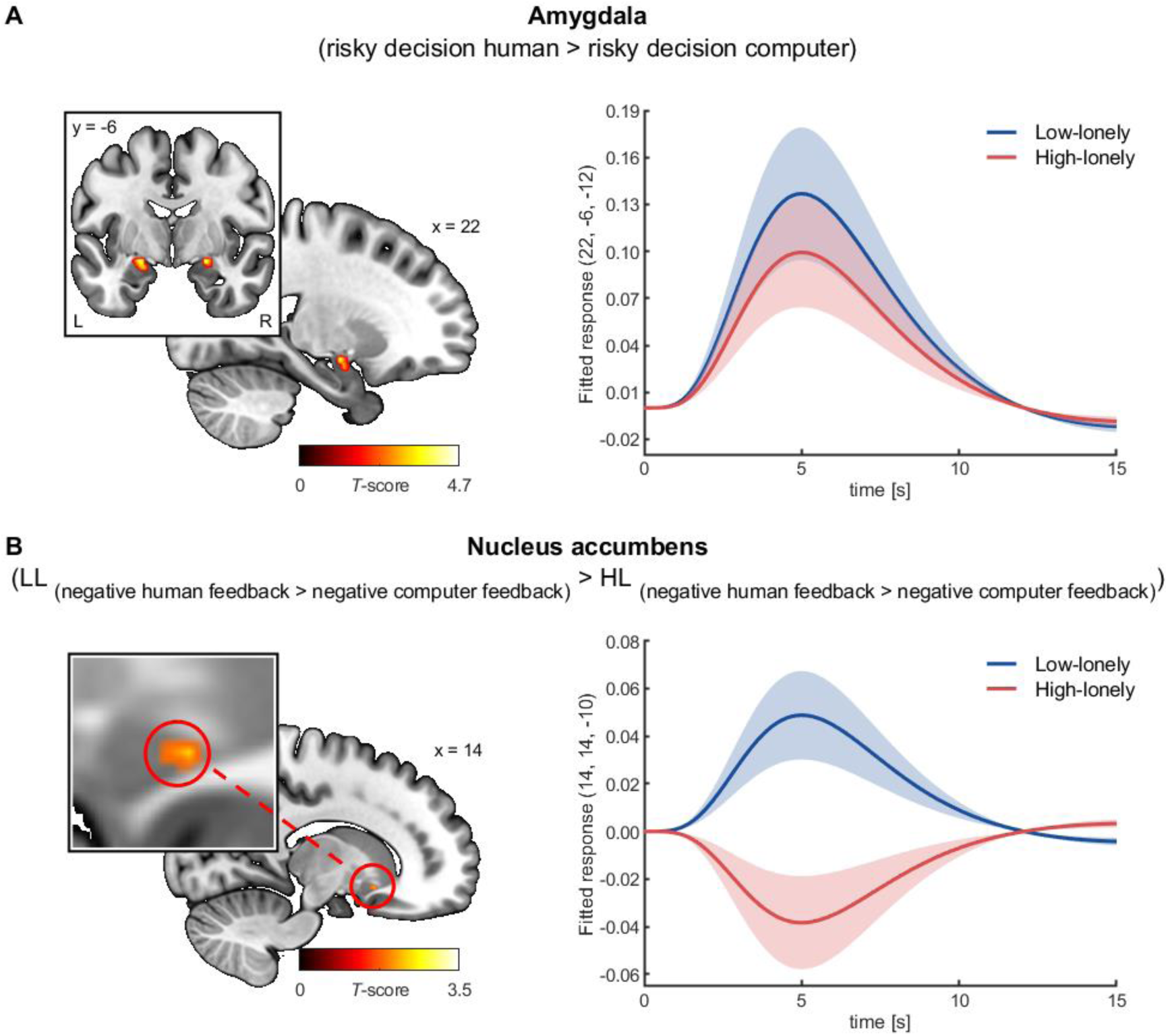
Neural activation during the social gambling task. (**A**) Amygdala activity was significantly enhanced during the decision phase of the social gambling task when participants chose the risky option with a human partner compared to the computer partner (right: 22, -6, - 12, t(73) = 4.58, P_FWE_ = 0.002; left: -22, -8, -12, t(73) = 4.23, P_FWE_ = 0.006). In line with the behavioral results, no group differences in neural activity were observed during the decision phase. (**B**) During the feedback stage, participants with high loneliness scores (HL) showed attenuated nucleus accumbens (NAcc) responses to negative feedback given by human partners compared to the computer partner. In contrast, NAcc reactivity to negative human feedback was enhanced compared to computer feedback in control participants (LL). Shaded areas show the standard error of the mean of the fitted responses based on the hemodynamic response function. For illustration purpose, clusters are shown with significance levels of P < 0.05 uncorrected. Abbreviations: L, left, R, right.

Importantly, however, amygdala activation during the decision or feedback stage did not significantly differ between HL and LL participants. Conversely, we observed significant differences in striatal responses to the feedback videos. HL participants showed significantly smaller NAcc responses to human (vs. computer) feedback videos than LL individuals (14, 14, -10, *t*(74) = 3.07, *P*_FWE_ = 0.02). Again, the group difference was specific for negative feedback videos (14, 14, -10, *t*(74) = 3.21, *P*_FWE_ = 0.01; see supplementary material and **Fig. 2B**), whereas no significant group effects were observed for responses to positive feedback videos. Post-hoc tests revealed increased NAcc responsiveness to negative human feedback compared to the computer feedback in LL participants (*t*(36) = 2.59, *P*_cor_ = 0.03, *d* = 0.53), while HL participants exhibited the opposite response pattern (*t*(38) = -1.96, *P*_cor_ = 0.12). No further group differences in brain activity were observed.

Exploratory gPPI analyses of the negative feedback condition with the NAcc serving as seed region indicated enhanced functional connectivity of the left NAcc with a cluster including the hippocampus in HL compared to LL participants (−14, -22, -14, *k* = 73, *t*(74) = 5.38, *P*_FWE_ = 0.049 on cluster level; see **Fig. 3A**). Again, post-hoc tests revealed an opposing pattern between groups with enhanced connectivity while receiving negative human (vs. computer) feedback in HL participants (*t*(38) = 3.06, *P*_cor_ = 0.01, *d* = 0.63) and reduced connectivity in LL participants (*t*(36) = -4.93, *P*_cor_ < 0.001, *d* = -1.15). Interestingly, NAcc-hippocampus connectivity not only correlated with NAcc responses to negative human feedback (contrasted with negative computer feedback: *r*(74) = -0.33, *P* = 0.004, i.e., increased connectivity was associated with reduced neural reactivity), but also with pleasantness ratings of negative feedback videos (*r*(74) = 0.23, *P* = 0.04, see **Fig. 3B**). The correlation between NAcc activity and negative feedback ratings was similar, but failed to reach significance (*r*(74) = -0.20, *P* = 0.09).

**Fig. 3.**
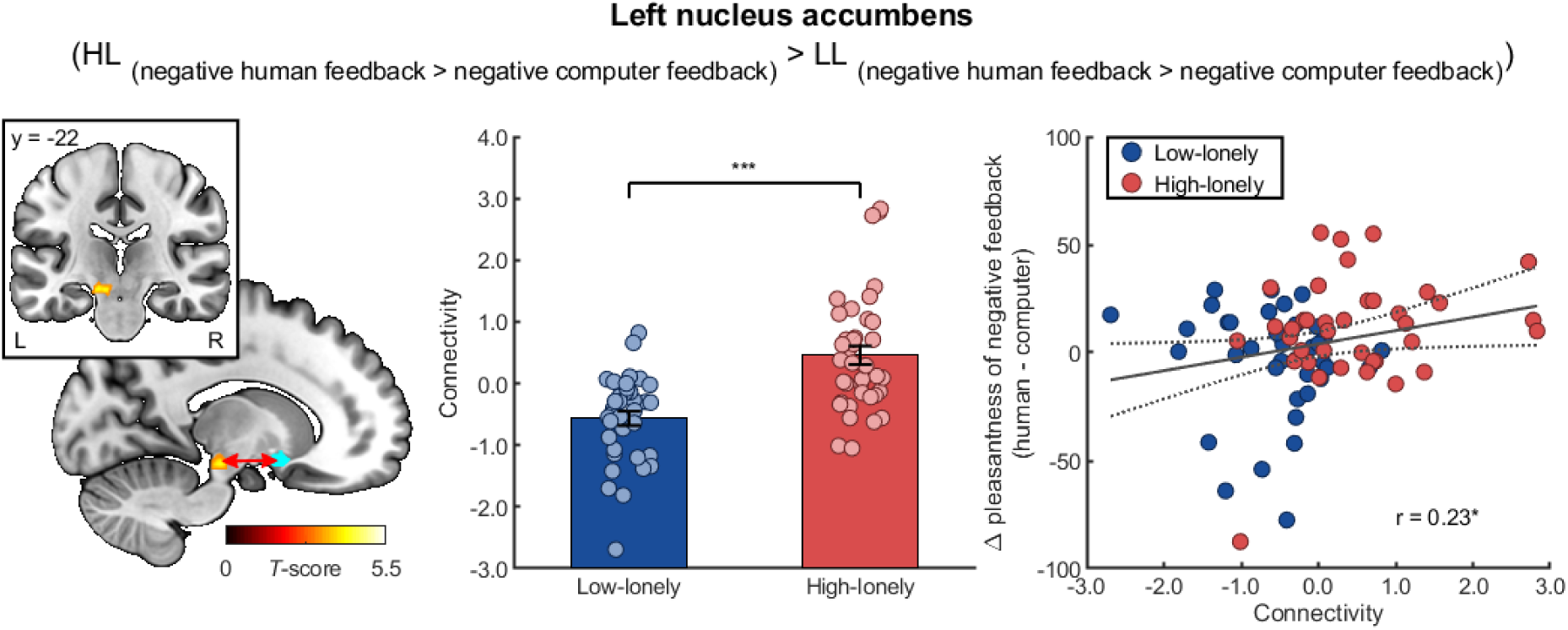
Functional connectivity during the social gambling task. Participants with high loneliness scores (HL) showed enhanced functional connectivity of the nucleus accumbens (blue sphere) with a cluster including the hippocampus while receiving negative human (vs. computer) feedback compared to control participants (LL). Functional connectivity positively correlated with the pleasantness ratings of the negative human feedback (compared to the negative computer feedback). The dashed line represents the 95%-confidence interval of the plotted regression line. Bars represent group means. Error bars indicate standard errors of the mean. Abbreviations: L, left, R, right. * P < 0.05, *** P < 0.001.

### Bayesian analyses and effects of confounding variables

Bayesian analyses revealed moderate evidence for the absence of group differences in variables that have previously been associated with social anxiety (cf. (31)), with our data being at least three times more likely under the null hypothesis (H0: no differences between groups) than under the alternative hypothesis (HL differ from LL participants in any direction). Specifically, Bayesian t-tests revealed moderate evidence that HL participants indeed did not differ from LL participants regarding the pleasantness ratings of positive human compared to computer feedback as our data were found to be almost four times more likely under the H0 than under the alternative hypothesis (Bayes factor (BF_10_) = 0.25, median effect size = 0.08, 95 % credible interval: [-0.32, 0.49]).

Likewise, Bayesian analyses revealed moderate evidence that groups showed equal reward-associated brain activity in response to positive human feedback (contrasted with positive computer feedback; left NAcc: BF_10_ = 0.25, median effect size = 0.07, 95 % credible interval: [-0.35, 0.49]; for the right NAcc the evidence is inconclusive: BF_10_ = 0.43, median effect size = 0.23, 95 % credible interval: [-0.19, 0.66]) and moderate evidence in favor of the H0 for amygdala reactivity to human feedback (contrasted with computer feedback; left: BF_10_ = 0.24, median effect size = -0.004, 95 % credible interval: [-0.42, 0.41]; right: BF_10_ = 0.24, median effect size ≈ 0.00, 95 % credible interval: [-0.42, 0.42]). The same pattern of results was observed for amygdala activation during the decision stage of the social gambling task as our data were up to four times more likely under the assumption of comparable activation between groups (H0) than under the alternative hypothesis (left amygdala activation for risky decisions with a human partner compared to a computer partner: BF_10_ = 0.24, median effect size = 0.03, 95 % credible interval: [-0.39, 0.45]; left amygdala activation for risky decisions with a human partner contrasted with safe decisions in trials with a human partner: BF_10_ = 0.33, median effect size = -0.17, 95 % credible interval: [-0.61, 0.25]; right: BF_10_ = 0.24, median effect size = -0.01, 95 % credible interval: [-0.43, 0.41]). For right amygdala activation, there was insufficient evidence to draw a conclusion for or against the hypothesis that groups exhibit equal responsiveness to risky decisions involving a human partner (contrasted with the computer; BF_10_ = 0.50, median effect size = 0.26, 95 % credible interval: [-0.16, 0.70]). However, descriptive analyses revealed an opposing response pattern in HL participants to what has been expected due to increased social anxiety symptoms: while LL participants showed slightly enhanced amygdala activation (mean parameter estimates ± SD: 0.25 ± 1.06), amygdala activation was reduced in HL participants (mean parameter estimates ± SD: -0.02 ± 0.68; cf. **Fig. 2A**). Likewise, no evidence for any of the hypotheses (null or alternative hypothesis) was observed for the subjective value of engaging in social situations (BF_10_ = 0.57, median effect size = -0.29, 95 % credible interval = [-0.74, 0.15]). Again, descriptive analyses revealed enhanced values of social engagement in HL compared to LL participants, which is contrary to the previously reported negative association with social anxiety (see inlay of **Fig. 1B** and cf. (31)).

Regarding the invested money during the virtual auction task, Bayesian analyses provided moderate evidence for comparable investments between groups to avoid negative social feedback (BF_10_ = 0.33, median effect size = 0.17, 95 % credible interval = [-0.23, 0.59]) or to receive positive social feedback (BF_10_ = 0.33, median effect size = 0.18, 95 % credible interval = [-0.23, 0.59]).

Mediation and moderation analyses indicated that none of the reported group effects was mediated or moderated by confounding psychiatric symptoms (see supplementary material). Moreover, the observed effects of loneliness (HL vs. LL) on NAcc responsiveness to negative human feedback remained significant after including the potential mediators in the regression models (*P*s < .01 for all direct effects of group after including the mediator). Likewise, loneliness effects on NAcc-hippocampal functional connectivity while receiving negative human feedback was found to be robust (all direct effects of group after including the respective mediator *P*s < .0001) and even increased after including social anxiety scores in the regression model (see supplementary material).

## Discussion

The current study sought to investigate shared and distinct behavioral and neural response patterns underlying social anxiety and loneliness. Our results revealed that a previously observed neurocircuitry underlying avoidance behavior in social anxiety (cf. (31)) could not be replicated in lonely individuals. HL participants differed from control participants neither in the subjective value of engaging in social situations nor in neural responses to social decision-making and positive social feedback. Conversely, HL participants showed altered responsiveness to negative social feedback evident in opposing behavioral response patterns and striatal brain activity and connectivity compared to control participants.

Our results thus indicate that loneliness might be more associated with altered emotional reactivity to social situations than with behavioral tendencies to withdraw from social interactions. Human and animal research have consistently shown that the amygdala is crucially involved in the processing of threat-related stimuli and hyperactivation of the amygdala is known as a core mechanism underlying anxiety disorders (30, 48). Moreover, amygdala habituation to threat-related stimuli and amygdala connectivity with prefrontal regions predict subsequent avoidance behavior (49-51). Likewise, we have previously found that amygdala activation during decisions in the social gambling task increases with social anxiety symptomatology and negatively correlates with the subjective value to engage in social situations (31). By contrast, the subjective value of engaging in a social situation did not differ between HL participants and controls and Bayesian analyses revealed evidence for comparable amygdala activation during the decision and feedback stages. In line with our findings, neuroanatomical correlates of social avoidance behavior were previously found to be unaffected by loneliness (52). This notion is consistent with etiological theories that highlight maladaptive social cognitions in the development and maintenance of loneliness (27, 53). Likewise, cognitive-behavioral interventions were found to be more effective in targeting social biases than social skill trainings (54, 55). There is preliminary evidence that established cognitive-behavioral treatments targeting social anxiety concurrently decrease feelings of loneliness and vice versa (56-60), but our findings of distinct behavioral and neural substrates suggest that loneliness-adjusted protocols might improve therapeutic outcomes.

Moreover, our results provide new insights into the neural pathways underlying loneliness. Unexpectedly, striatal activity during negative social feedback was reduced while pleasantness ratings were increased in HL participants. Notably, activation of the NAcc is associated with goal-directed approach and avoidance behavior and involved in avoiding social punishment (61-63). As HL participants rated the negative social feedback videos as more pleasant than the control participants, reduced NAcc responses to negative social feedback might thus reflect reduced tendencies to avoid this negative social feedback. Furthermore, the enhanced functional coupling of the NAcc with a hippocampal cluster that correlated with individual pleasantness ratings is in line with the involvement of this neural circuit in hedonic processing (64) and might reflect the rewarding experience of a social feedback for socially deprived individuals (65). Nevertheless, we have recently found a compromised neural integration of social information in HL participants evident in various brain regions including the NAcc (37). Furthermore, loneliness has been associated with a reduced recognition of negative vocal expressions (66). Thus, the reduced NAcc activity might also reflect diminished differentiation between positive and negative feedback, resulting in a dysregulated reward system responsiveness to negative social stimuli as observed for the NAcc-hippocampus connectivity. However, inference about cognitive processes from neural activation should always be drawn with restraint (67) and results regarding biased emotion recognition in loneliness are inconclusive (68). Future studies are warranted to further investigate the impact of loneliness on the processing of negative social feedback.

Interestingly, differences between HL and control participants were restricted to behavioral and neural responses to negative social feedback, whereas Bayesian analyses revealed evidence for a comparable responsiveness to positive social feedback between groups. Conversely, social anxiety has been consistently found to affect the processing of social rewards (31-34). Previous studies point to various negative effects of loneliness on the processing of positive social interactions (37, 69, 70), but findings about the association between loneliness and NAcc reactivity to positive social stimuli are mixed. The involvement of the NAcc in loneliness might be context-dependent, with feelings of social isolation promoting the hedonic experience of positive social stimuli in an acute stage (65), which may be different from chronic loneliness. Similarly, lonely individuals might experience a social stimulus as more rewarding only if the stimulus is already familiar (e.g. a romantic partner and not a stranger (71)). Along these lines, a recent study found no relationship of loneliness with striatal responsiveness to pictures depicting strangers during positive social interactions (72). Nevertheless, in our task design positive feedback was always coupled with monetary gains. Thus, differences regarding positive social feedback might have been obfuscated by the rewarding experience of earning money as evident in enhanced striatal responsiveness to positive feedback, irrespective of the partner providing the feedback. Both external (e.g., passive viewing of positive social interactions vs. being involved in a positive social interaction) and internal factors (e.g., state vs. chronic feelings of social isolation) may influence the association of loneliness with social reward processing.

Moreover, given the quasi-experimental, cross-sectional design of our study, our findings do not allow casual inferences about the relationship of loneliness and social feedback processing. In addition, moderation and mediation analyses indicate that the observed associations with loneliness were not driven by psychiatric symptoms that were also more pronounced in HL individuals. However, our study specifically focused on high-lonely healthy individuals who may represent a resilient subsample of the population because they did not develop acute psychiatric disorders. Thus, clinical studies with psychiatric patients are warranted to uncover the direction of the observed associative relationships and to further disentangle shared and distinct mechanisms underlying loneliness and psychopathology.

Collectively, the current results suggest that loneliness and social anxiety are distinct constructs with specific behavioral and neural substrates. Along these lines, interventions targeting loneliness-specific cognitive biases may be more effective in reducing loneliness than cognitive behavioral therapies focused on reducing avoidance behavior.

## Data Availability

All data used in this study are openly available (https://osf.io/p6jxk/ and https://neurovault.org/collections/VNYRMORR/).

https://neurovault.org/collections/VNYRMORR/

## Acknowledgement

The authors thank Alexandra Goertzen-Patin for proofreading the manuscript.

## Author Contributions

J.L., J.S., and D.S. designed the experiment; J.L., T.E., and E.K. conducted the experiments; J.L. and D.S. analyzed the data. All authors wrote the manuscript. All authors read and approved the manuscript in its current version.

