## Supplementary Information for "Behavioral and neural dissociation of social anxiety and loneliness"

### Supplementary Materials and Methods

#### Details of the pre-stratification process and study design

Loneliness scores were assessed via an online survey that included the revised version of the UCLA loneliness scale (UCLA-L, (1)). Participants with UCLA-L scores of 50 or above (i.e., at least one standard deviation above the mean score of students, cf. (1)) were assigned to the high-lonely (HL) group, while participants with scores of 25 or below (i.e., at least one standard deviation below the mean score of students) were assigned to the control (low-lonely, LL) group. Out of a sample of 3,678 adults who completed the UCLA loneliness scale, 91 individuals were assigned to either the HL or the LL group, agreed to participate and were invited to a screening session prior to the test session to assess eligibility of enrollment. Nine participants had to be excluded after the screening session since they did not fulfill the inclusion criteria (aged 18-65, no current physical or psychiatric disorder as assessed via self-disclosure and by the Mini-International Neuropsychiatric Interview (2), no psychotherapy, no current psychotropic medication, no illicit drug use in the previous four weeks, right-handed, eligibility for magnetic resonance imaging scanning), resulting in the final sample of 42 HL (21 female; mean UCLA-L score ± standard deviation (SD): 57.05 ± 5.41) and 40 LL (20 female; mean UCLA-L score ± SD: 23.78 ± 1.25) participants. Following the screening session, participants completed the virtual auction task. The social gambling task was completed during a separate test session and repeated during functional magnetic resonance imaging (fMRI) on the same day. Data collection was completed before the start of the COVID-19 pandemic.

### Power Analysis

The sample size was based on an a-priori power analysis (cf. (3)). To determine an effect size that could be expected for loneliness regarding altered neural processing of social stimuli, we referred to (4). The analysis using G*Power 3 (5) indicated that at least 71 participants were needed to reliably replicate the reported loneliness effect on ventral striatum/amygdala activity with a power of 0.99 ($\alpha = 0.05$). To account for possible missing data and drop-outs, we planned to test at least 80 participants.

### Details of the virtual auction task

In each trial, a picture of one of six actors indicated which feedback video was being auctioned. One trial was chosen randomly after the completion of the six trials in one condition (i.e., positive or negative feedback) and the invested money was compared to the invested money by the computer. The player (participant or computer) who invested more money won the auction and kept the remaining money (1 € minus the invested money). If the participant won during the positive feedback condition, a positive social feedback video (expressing admiration) was presented after winning the auction, while no video was presented during the negative feedback condition. If the participant lost, a negative social feedback video (expressing condescension) was presented during the negative feedback condition and no video was shown in the positive feedback condition. If the participant lost, the participant kept 1 €, irrespective of the invested money. All pictures and videos were taken from a validated database (6). The feedback videos were repeated until the participants pressed any key. Participants received the remaining money of the randomly chosen trials.

### Details of the social gambling task

For further details of the implementation of the social gambling task, see (7). Each trial of the social gambling task consisted of a decision and a feedback stage. During the decision stage, participants could decide between the risky (i.e., a dice game with the chance to win 3 €) and the safe option (i.e., a fixed payoff ranging from 0 to 3 € in steps of 50 cents) with no imposed time limit. The task contained a human (indicated by the name and picture of one of four partners) and a computer control condition (indicated by a picture of a computer). If participants chose the risky option in the human condition, either a positive or a negative social feedback video was shown during the feedback stage, dependent on the outcome of the trial. The same actors and videos as included in the virtual auction task were used. Each human partner (four different partners) was paired twice with each possible amount of money (0 to 3 € in steps of 50 cents) offered as alternative for the risky option, resulting in 56 trials. Likewise, participants completed 56 trials of the control condition. After completing the behavioral task, participants were asked to rate the pleasantness of each positive and negative feedback video on a visual analogue scale (VAS; ranging from 0 “not pleasant at all” to 100 “very pleasant”). For each participant, the individual certainty equivalents (CE20, CE50, CE80) were estimated by fitting a cumulative Gaussian function to each participant’s choice probabilities observed for the payoffs offered in the safe option and defined as the payoff a participant would have to receive to attain a 20 %, 50 %, and 80 % chance of choosing the safe option.

The task was repeated during fMRI with the following adjustments: the partner for each trial (one of four human partners or the computer) was chosen randomly and indicated by the name of the partner or the word “computer”. Furthermore, the fixed payoff offered in the safe option varied randomly between the three individually determined values CE20, CE50, and CE80. Participants responded with their index fingers using an MRI-compatible response grip system (NordicNeuroLab AS, Bergen, Norway). The position of the risky option (left or right on the screen) was counterbalanced across trials. All human partners were presented in combination with each of the three CE values twice, resulting in 24 human trials and 24 computer trials per run. The feedback video was presented for 2.6 seconds. The temporal intervals between the decision and outcome stages and the inter-stimulus intervals between trials varied from 2 to 11 seconds with a descending probability. All participants completed two runs. Participants received the obtained money from one randomly chosen trial per run.

**Behavioral data analysis**

We calculated mixed ANOVAs with the estimated CE50 values, the proportion of safe decisions during the behavioral and the fMRI task, and the pleasantness ratings of the feedback videos as dependent variables. For all analyses, group (HL vs. LL) served as between-subject factor and the partner condition (human vs. computer) was included as within-subject factor. Offered payoffs as safe option were further included as within-subject factor for the behavioral task (0 € to 3 €) and the fMRI task (CE20, CE50, CE80) to analyze the proportion of safe decisions, whereas the analysis of the pleasantness ratings of the feedback videos included the additional within-subject factor feedback valence (positive vs. negative feedback). For the analysis of the virtual auction task, effects of the valence (positive vs. negative video) and group were included as within- and between-subject factors, respectively, in a mixed ANOVA with invested money serving as dependent variable. Greenhouse-Geisser corrections were applied in cases of violated assumptions of sphericity as tested by Mauchly's test. All post-hoc t-tests to disentangle interactions were Bonferroni-corrected ($P_{cor}$). Pearson's product-moment correlations were calculated to investigate the relationship of observed behavioral group effects with neural group effects.

### fMRI data acquisition and preprocessing

All fMRI data were acquired using a 3T Siemens TRIO MRI system (Siemens AG, Erlangen, Germany) with a Siemens 32-channel head coil. Functional data of the social gambling task were acquired using a T2*-weighted echoplanar (EPI) sequence with a repetition time (TR) of 2500 ms, an echo time (TE) of 30 ms, ascending slicing, a matrix size of 96 x 96, 37 axial slices with a voxel size of 2 x 2 x 3 $mm^3$ and a slice thickness of 3.0 mm, a distance factor of 10 %, a field of view (FoV) of 192 x 192 $mm^2$, and a flip angle of 90°. High-resolution T1-weighted structural images were collected on the same scanner (TR = 1660 ms, TE = 2.54 ms, matrix size: 256 x 256, voxel size: 0.8 x 0.8 x 0.8 $mm^3$, slice thickness = 0.8 mm, FoV = 256 x 256 $mm^2$, flip angle = 9°, 208 sagittal slices). To control for inhomogeneity of the magnetic field, fieldmaps were obtained for the T2*-weighted EPI sequence (TR = 392 ms, TE [1] = 4.92, TE [2] = 7.38, matrix size: 64 x 64, voxel size: 3 x 3 x 3, slice thickness = 3.0 mm, distance factor = 10 %, FoV = 192 x 192 $mm^2$, flip angle 60°, 37 axial slices). For preprocessing, standard procedures of SPM12 (Wellcome Trust Center for Neuroimaging, London, UK; http://www.fil.ion.ucl.ac.uk/spm) implemented in Matlab (The MathWorks Inc., Natick, MA) were used. The first five volumes of each functional time series were removed to allow for T1 signal equilibration before affine registration was used to correct for head movements between scans. Images were initially realigned to the first image of the time series and then re-realigned to the mean of all images. For unwarping, the voxel displacement map (VDM file) was applied to the EPI time series to correct for signal distortion based on B0-field inhomogeneity. Normalization parameters as determined by segmentation and non-linear warping of the structural scan to reference tissue probability maps in Montreal Neurological Institute (MNI) space were applied to all functional images. All images were resampled at 2 x 2 x 2 $mm^3$ voxel space and spatially smoothed by using a 6-mm full width half maximum (FWHM) Gaussian kernel. A high-pass filter with a cut-off period of 128 s was used to detrend raw time series.

### fMRI data analysis

To analyze the fMRI data, we used a two-stage approach as implemented in SPM12. On the first level, data were modeled using a fixed-effects model. Onsets and durations of eight conditions (‘risky decision computer’, ‘safe decision computer’, ‘risky decision human’, ‘safe decision human’, ‘positive computer feedback, ‘negative computer feedback’, ‘positive human feedback’, ‘negative human feedback’) were modeled by a stick function convolved with a hemodynamic response function (HRF). Although individual CE values were used during the fMRI task to equalize the number of trials of each condition between both runs, the decisions of the participants and thereby the resulting number of trials of one condition still differed between runs to varying degrees. We thus decided to concatenate time series of both runs (cf. (8)). Baseline regressors were added for each run, and the high-pass filter and temporal non-sphericity estimates were adjusted separately for each run. The six movement parameters were included in the design matrix as regressors of no interest. Within-subject contrasts of interest were calculated on the first level and entered to a random-effects model on the second level. For task validation, one-sample t-tests were calculated across groups (i.e., decision human > decision computer, risky decision human > risky decision computer, safe decision human > safe decision computer, human feedback > computer feedback, positive feedback > negative feedback). Furthermore, whole-brain task effects (e.g., decision human > decision computer) were analyzed across groups after applying an initial cluster-forming height threshold of $P < 0.001$. Five participants were excluded from fMRI analyses due to excessive head movement (> 4 mm/° in any direction; $n = 2$), anatomical abnormalities ($n = 1$), technical issues ($n = 1$), or incomplete data ($n = 1$). Furthermore, three participants were excluded from analyses of the decision stage as they always chose the risky option for at least one of the partners, while one participant was excluded from analyses of the feedback stage because no positive human feedback was shown during both runs.

### Multivariate pattern analysis (MVPA)

For the decoding analysis, we used non-normalized and unsmoothed data of each participant and included the same conditions and regressors as outlined above in the single-subject fixed-effects models separately for both runs. The participants' decisions (risky or safe decision) were used as independent variables and parameter estimates of the corresponding first level regressors were used as features. Using the default parameters of the Decoding Toolbox (9), we ran a classification searchlight analysis with a 9-mm searchlight radius and trained a support vector machine classifier (LIBSVM) on the data of one run to decode the decision to play or to choose the safe option. The decoding accuracy was tested on the data of the other run and the resulting individual accuracy maps minus chance (chance = 50 % accuracy) were normalized to MNI space and smoothed using a 6-mm FWHM Gaussian kernel. Accuracy maps were then entered to a random-effect model on the second level and tested against 0 by calculating a one-sample t-test across groups. Familywise error (FWE) correction was applied based on the size of the anatomically defined amygdala (cf. (7)).

### Functional connectivity analyses

Preprocessing for the exploratory functional connectivity analyses additionally included a denoising pipeline. Following the recommendations of the CONN toolbox 19.b (www.nitrc.org/projects/conn, RRID:SCR_009550), outlier scans were detected by the integrated artefact detection toolbox-based identification using conservative settings (i.e., thresholds of 0.5 mm frame wise displacement and 3 SD above global BOLD signal changes were used) and treated as regressors of no interest in the following analyses . The default denoising pipeline implemented a linear regression of confounding effects of the first five principal noise components from white matter and cerebrospinal fluid template masks, 12 motion parameters, scrubbing, and constant task-related effects. A high-pass filter of 0.008 Hz was applied to minimize the effects of physiological and motion related noise. Regions associated with group effects (amygdala or NAcc) served as seed regions in a seed-to-voxel analysis. The interaction terms of the psychological (task conditions convolved with a canonical HRF) and the physiological factor (blood oxygenation level dependent signal) were computed for each participant on the first level. The relative measure of connectivity compared to the implicit baseline was calculated by using bivariate regression measures. Connectivity was compared between groups on the second level by using mixed analyses of variance (ANOVA).

#### Mediation and moderation analyses

To examine the influence of possible confounding variables on significant group effects (i.e., depressive symptomatology assessed by the Beck's Depression Inventory II, BDI (10), childhood maltreatment assessed by the Childhood Trauma Questionnaire, CTQ (11), and social anxiety measured by the Liebowitz Social Anxiety Scale, LSAS (12)), we calculated mediation and moderation analyses using the PROCESS macro v3.4 for SPSS (13). BDI, CTQ, and LSAS scores were used as mediator and moderator variables and group as predictor variable. For mediation analyses, 10,000 bootstrap samples were used. Variables were mean-centered before calculating moderation analyses.

#### Bayesian analyses

For all hypothesized differences between HL and LL participants that could not be confirmed by classical inference analyses, Bayesian t-tests were conducted to quantify the evidence for the null hypotheses (i.e., HL participants do not differ from LL participants) using the default settings for two-tailed independent t-tests implemented in JASP (14). Specifically, group differences in the subjective value of engaging in social situations during the social gambling task (i.e., the individual

CE50 for human partners minus CE50 for the computer partner) and pleasantness ratings of positive human feedback (minus the ratings of positive computer feedback) were re-analyzed by calculating Bayesian t-tests. Moreover, as we expected HL participants to differ from LL participants regarding amygdala responsiveness to risky decisions involving a human partner, parameter estimates during the decision stage of the anatomical amygdala were averaged across all voxels and re-analyzed to quantify evidence that neural activation is equal between groups for the following contrasts of interest: risky decision human > risky decision computer and risky decision human > safe decision human. Likewise, parameter estimates of activation during the feedback stage were extracted from the amygdala to re-analyze responsiveness to human feedback (compared to computer feedback). To re-analyze reward-associated brain activity in response to positive human feedback (compared to computer feedback), parameter estimates were extracted from the NAcc.

### Supplementary Results

#### Additional behavioral results

The proportion of safe decisions in the behavioral social gambling task significantly increased with higher payoffs offered as safe alternative to the risky gambling decision across groups (main effect of offered payoff: $F(2.95,236.14) = 183.77$, $P < 0.001$, $\eta_p^2 = 0.70$; see **Fig. 1A**) and was highest for an offered payoff of 3 € (mean proportion of safe decisions ± SD for an offered payoff of 0 €: 8.16 ± 17.06 %; 0.5 €: 8.38 ± 16.44 %; 1 €: 19.36 ± 28.44 %; 1.5 €: 37.96 ± 36.12 %; 2 €: 76.98 ± 30.70 %; 2.5 €: 84.98 ± 25.85 %; 3€: 88.11 ± 23.48 %). Post-hoc tests comparing adjacent payoffs revealed significant differences for all comparisons except for the likelihood of safe decisions for 0 € versus 0.5 € ($t(81) = 0.14$, $P_{cor} > 0.05$) and 2.5 € versus 3 € payoffs ($t(81) = 1.51$, $P_{cor} > 0.05$; all other $t$s > 3.46, $P_{cor} < 0.01$, $d$s > 0.27). Likewise, the proportion of safe decisions differed between all three payoffs offered during the fMRI implementation of the task (main effect of offered payoff: $F(2,158) = 185.43$, $P < 0.001$, $\eta_p^2 = 0.70$; post-hoc comparisons: CE20 vs. CE50: $t(80) = 8.27$, $P_{cor} < 0.001$, $d = 1.08$; CE50 vs. CE80: $t(80) = 11.02$, $P_{cor} < 0.001$, $d = 1.44$). Moreover, in the behavioral task, a significant interaction of partner and payoffs revealed less safe decisions for the human partner compared to the computer when the offered payoff was low, while this effect was reversed when the offered payoff was high ($F(3.63,290.72) = 5.74$, $P < 0.001$, $\eta_p^2 = 0.07$; post-hoc comparisons of the proportion of safe decisions between the human and computer partner for an offered payoff of 0 €: $t(81) = -4.13$, $P_{cor} < 0.001$, $d = -0.33$; 0.5 €: $t(81) = -3.57$, $P_{cor} = 0.004$, $d = -0.29$; 1 €: $t(81) = -3.50$, $P_{cor} = 0.005$, $d = -0.23$; 3 €: $t(81) = 2.84$, $P_{cor} = 0.04$, $d = 0.26$; all other $t$s > -0.09 and < 1.06, $P_{cor} > 0.05$). Importantly, as individual payoffs were calculated for the fMRI task separately for human and computer partners to equalize the ratio of risky and safe decisions, the likelihood of safe decisions during fMRI differed neither between partners nor between groups (HL vs. LL) (all main effects or interactions of the partner condition or group *F*s < 1.48, *P*s > 0.05).

As intended, positive feedback videos were rated as more pleasant than negative ones (main effect of feedback valence: $F(1,80) = 174.73$, $P < 0.001$, $\eta_p^2 = 0.69$). In addition to the reported three-way interaction (see main results), a significant interaction of partner and feedback valence was observed ($F(1,80) = 5.45$, $P = 0.02$, $\eta_p^2 = 0.06$), indicating that ratings differed more between human and computer partners in the positive feedback condition than in the negative one. No further significant effects were observed for the social gambling task or the virtual auction task.

**Detailed results of mediation and moderation analyses**

Groups differed significantly regarding psychiatric symptoms (cf. (3)). HL participants reported more depressive symptoms ($t(50.89) = 4.15$, $P < 0.001$, $d = 0.92$; mean BDI score ± SD in HL: 6.62 ± 6.76; LL: 2.03 ± 2.31) and more severe childhood maltreatment ($t(80) = 2.38$, $P = 0.02$, $d = 0.53$; mean CTQ score ± SD in HL: 38.86 ± 10.28; LL: 31.90 ± 15.76; for social anxiety symptomatology, see main text). We thus tested whether the observed explorative group effects were mediated or moderated by depressive or social anxiety symptomatology and childhood maltreatment. Our results revealed that none of the reported group effects was significantly mediated or moderated by psychiatric symptoms (the 95 % confidence interval (CI) of all tested indirect effects included zero and all interaction effects of group with the potential moderator $P > 0.05$), except for NAcc responsivity to the negative human feedback (contrasted with the negative computer feedback). Analyses showed a significant suppressor effect of social anxiety on the relationship between group and NAcc responses (indirect effect of group on NAcc activity via social anxiety: $\beta = 0.14$, SE = 0.11, 95 % CI: 0.004 to 0.40). Thus, the absolute height of the group effect even increased after including social anxiety as mediator (effect of group without taking social anxiety into account: β = -0.69, SE = 0.22, 95 % CI: -1.12 to -0.26; with social anxiety as mediator: β = -0.83, SE = 0.23, 95 % CI: -1.28 to -0.38).

### Supplementary Tables

#### Table S1. Whole-brain findings across groups

| Region | Right/left | Cluster size (voxel) | Peak *T* | MNI coordinates | | |
|---|---|---|---|---|---|---|
| | | | | x | y | z |
| **Decision human > decision computer** | | | | | | |
| Medial orbitofrontal gyri | bil. | 351 | 6.28 | 2 | 44 | -14 |
| Precuneus | bil. | 800 | 6.04 | 4 | -56 | 28 |
| **Risky decision human > risky decision computer** | | | | | | |
| Superior temporal gyrus | R | 448 | 7.60 | 48 | -40 | 10 |
| Precuneus | bil. | 496 | 6.64 | 6 | -56 | 28 |
| Medial orbitofrontal gyri | bil. | 328 | 5.79 | 2 | 42 | -14 |
| Inferior frontal gyrus, triangularis | R | 315 | 5.49 | 42 | 16 | 22 |
| **Human feedback > computer feedback** | | | | | | |
| Middle temporal gyrus | R | 6,837 | 12.07 | 54 | -40 | 8 |
| Calcarine fissure | R | 141 | 12.01 | 22 | -94 | -2 |
| Amygdala | L | 3,273 | 9.66 | -22 | -8 | -12 |
| Fusiform gyrus | R | 361 | 9.29 | 40 | -48 | -16 |
| Fusiform gyrus | L | 296 | 8.44 | -38 | -48 | -20 |
| Middle occipital gyrus | L | 32 | 7.65 | -20 | -94 | -2 |
| Gyri rectus | bil. | 295 | 6.54 | 6 | 38 | -16 |
| Inferior occipital gyrus | R | 42 | 5.29 | 44 | -76 | -6 |
| **Positive feedback > negative feedback** | | | | | | |
| Inferior occipital gyrus | R | 341 | 8.32 | 26 | -92 | -2 |
| Caudate nuclei | bil. | 2,792 | 8.10 | 8 | 10 | -2 |
| Middle cingulate gyri | bil. | 2,897 | 6.80 | -2 | -34 | 34 |
| Inferior occipital gyrus | L | 101 | 6.63 | -28 | -88 | -6 |
| Angular gyrus | L | 3,721 | 6.15 | -40 | -66 | 46 |
| Middle frontal gyrus | L | 2,771 | 6.11 | -30 | 16 | 52 |
| Precentral gyrus | R | 2,059 | 5.62 | 36 | -28 | 62 |
| Superior frontal gyrus | R | 722 | 5.59 | 20 | 34 | 48 |

| | | | | | | |
|---|---|---|---|---|---|---|
| Inferior orbitofrontal gyrus | L | 55 | 5.53 | -26 | 30 | -16 |
| Fusiform gyrus | L | 229 | 5.43 | -26 | -46 | -18 |

*Notes.* Cluster-sizes are based on the initial cluster-forming height threshold of $P < 0.001$. Peak $T$ and MNI coordinates are listed for FWE-corrected $P$s < 0.05 on peak level. No cluster survived the FWE-correction on the peak level for the safe decision human > safe decision computer contrast. For the positive feedback > negative feedback contrast, the nucleus accumbens is included in the caudate nuclei cluster. Abbreviations: bil., bilateral; L, left; MNI, Montreal Neurological Institute; R, right.
